# Modeling disease progression and placebo response in Huntington’s Disease: Insights from Enroll-HD and GENERATION HD1 cohorts

**DOI:** 10.1101/2024.02.15.24302445

**Authors:** Marcelo Boareto, Yumi Yamamoto, Jeffrey D. Long, Cristina Sampaio, Peter McColgan, Cheikh Diack, Patricia Sanwald Ducray

## Abstract

**Objective:** The objective of this study was to quantify longitudinal changes in Unified Huntington’s Disease Rating Scale (UHDRS) clinical scores and evaluate their susceptibility to placebo response, improving our understanding of disease progression and the ability to optimize clinical trials in Huntington’s disease.

**Methods:** We utilized data from the Enroll-HD and GENERATION HD1 placebo arm cohorts to model the natural history of the disease and placebo response for functional, motor and cognitive clinical scores. Baseline patient characteristics available in both cohorts were analyzed as potential predictors of progression rate.

**Results:** We identified distinct trajectories for each clinical measurement throughout the course of the disease, with baseline characteristics serving as strong predictors of progression rate. Interestingly, we observed a weak correlation between progression rates among different endpoints, highlighting the need for careful selection of endpoints, as they may not have the same relevance for different patient populations.

To quantify placebo response, we compared disease progression in the largest Huntington’s clinical trial to date (GENERATION HD1 placebo arm) to the Enroll-HD cohort. We found a strong initial improvement in motor and cognitive scores in GENERATION HD1, which remained constant during the dosing period of the trial. We show that this initial improvement in motor and cognitive scores can be quantified, enabling the simulation of changes in endpoints in a placebo arm for distinct subpopulations.

**Interpretation:** Our model successfully captures essential aspects of disease progression and placebo response in Huntington’s disease, providing valuable guidance for the design and analysis of upcoming clinical trials.

## Introduction

Disease progression in Huntington’s disease (HD) is complex and multifaceted, characterized by psychiatric disturbance, motor dysfunction, cognitive and functional impairment. These changes can be assessed using various clinical scores which are part of the Unified Huntington’s Disease Rating Scale (UHDRS), such as the Total Functional Capacity (TFC), Total Motor Score (TMS), Symbol Digit Modalities Test (SDMT), and Stroop Word Reading (SWR). Moreover, these scores can be integrated into a composite UHDRS score (cUHDRS, Schobel et al, 2017).

Currently, there is no approved disease-modifying therapy for HD. As with other rare diseases, numerous challenges exist in developing novel treatments, and there is a pressing need to optimize clinical trials in HD. This can be accomplished by more precisely identifying patients who are likely to exhibit disease progression during the trial period and determining the most appropriate endpoint for a specific patient population. Furthermore, sample size calculations typically rely on real-world data, and the impact of placebo response on the ability to differentiate a treatment effect from a placebo effect is not well understood. Therefore, a deeper understanding and quantification of longitudinal changes in clinical scores and placebo response are important steps in improving the design and analysis of clinical trials for HD.

Mathematical and computational methods have played an important role in advancing our understanding of disease progression in HD. One primary focus of these approaches has been predicting motor onset. The expansion of the cytosine-adenine-guanine (CAG) repeat lengthens the encoded polyglutamine segment of the huntingtin protein, and has been shown to be a strong predictor of age of motor onset (Andrew et al, 1993, Lee et al, 2022), with various groups characterizing a mathematical relationship between CAG repeat length and the age of motor onset that accounts for over half of its variance (Langbehn et al., 2010, Zhang et al 2011, Warner et al, 2022). Numerous studies have extended the prediction of motor onset beyond CAG repeat length by incorporating longitudinal clinical and imaging data (Paulsen et al., 2014), and also by deriving a prognostic measure for predicting disease progression and risk of motor diagnosis in premanifest individuals (Long et al 2017, Langbehn et al, 2022). These studies have significantly improved our prognostic capability concerning age of onset in HD, a relevant achievement since identifying patients who are likely nearing disease onset is essential for implementing clinical interventions early in the course of the disease, when preventing neuronal death and preserving function are most likely to happen.

In the past years, there has been a growing emphasis on expanding the assessment of HD beyond motor measures. Numerous studies have incorporated biological, clinical, and functional evaluations, including the proposal of a new staging system: the Huntington’s Disease Integrated Staging System (HD-ISS) (Tabrizi et al, 2022). Disease progression models taking into account various aspects of the disease have been proposed. Disease progression in HD has been suggested to happen as a series of states with increasing severity (Sun et al., 2019; Mohan et al., 2021). Predictive features of disease progression have been identified (Sun et al, 2020; Ghazaleh et al, 2021; Ko et al., 2022), and different disease trajectories have been categorized (Ko et al., 2022). Correlations between changes in features extracted from brain imaging or molecular biomarkers and clinical progression have been established (Tabrizi et al, 2012, Abeyasinghe et al, 2021, Rodrigues et al, 2020, Byrne et al, 2017). These efforts, along with the availability of large natural history studies such as Enroll-HD (Landwehrmeyer et al, 2017), have substantially improved our understanding of disease progression in HD and the key characteristics underlying individual variability in disease progression.

In this study, we developed a mathematical framework to investigate and quantify the natural history progression and placebo response in HD. Building on previous studies, we use data from Enroll-HD to infer optimal disease trajectories, quantify patient variability in progression rate, and predict progression based on patient characteristics. To gain a deeper understanding of placebo response in HD, we compared the progression in Enroll-HD (annual visits) with data from short-interval follow-up placebo data (GENERATION HD1, McColgan et al, 2023). Through this comparison, we were able to characterize and quantify placebo response, i.e., the variations in disease progression between natural history and the placebo arm of a pivotal trial. Lastly, we show that by quantifying both natural history progression and placebo response, we can simulate placebo arm outcomes for patient populations with different inclusion criteria.

## Methods

### Study Cohorts

Natural history data used in this work were generously provided by the participants in the Enroll-HD study and made available by CHDI Foundation, Inc. Enroll-HD is a global clinical research platform designed to facilitate clinical research in Huntington’s disease. We utilized data from Enroll-HD (NCT01574053) version PDS 5. We selected individuals with an HD-ISS Stage > 1 and at least one follow-up visit, which were conducted annually, resulting in a total of 6820 individuals selected. While participants were followed up to 9 visits, it is important to note that the majority of individuals had fewer than 3 follow-up visits.

To evaluate potential placebo response in HD, we used data from GENERATION HD1 (NCT03761849, McColgan et al, 2023). This double-blind Phase III study was designed to assess the efficacy and safety of tominersen in HD patients. The study enrolled 791 adults, with inclusion criteria including age between 25-65 years, Diagnostic Confidence Level (DCL) = 4, Independence Scale (IS) ≥70, CAG ≥ 36, CAG-age product (CAP) > 400, Body Mass Index (BMI) between 16-32 kg/m2, and weight exceeding 40 kg. Participants were evenly distributed into three groups: a placebo group and two active groups that received tominersen at two different dosing regimens. Patient monitoring occurred every 8 weeks. Based on an overall benefit–risk assessment by an independent data monitoring committee, dosing was halted after the majority of patients had completed 69 weeks of the trial. Patients continued to be followed until the 101st week. For the purpose of this analysis, only participants from the placebo group (n=260) were included.

For a comparative analysis of the baseline characteristics distribution between both cohorts, please refer to Figure Suppl 1 and McColgan et al. 2023.

### Mathematical model

In our approach, we begin by estimating the disease trajectories for each individual clinical score (TFC, TMS, SDMT, and SWR) throughout the course of the disease. We then determine the variability in the rate of progression and identify which covariates are predictive of this progression rate.

Our analysis involved specifying a hierarchical Bayesian model. This model incorporated fixed effects for the model structure and covariate coefficients, as well as random effects to account for subject-level variability. For more detailed information, please refer to the supplementary information.

To implement our model, we utilized the probabilistic programming language Turing.jl (Ge et al, 2018), which enables us to perform full Bayesian inference. Specifically, we employed Hamiltonian Monte Carlo (HMC), a Markov chain Monte Carlo (MCMC) method, to generate samples from the posterior distribution.

#### Covariate Selection

Patient characteristics at baseline were evaluated as potential predictive covariates of the progression rate. The following covariates were included: baseline levels of clinical scores or disease stage (TFC, TMS, SDMT, SWR, IS, HDISS), patient demographics (sex, height, weight, BMI, education level, age), co-medication (such as use of tetrabenazine, antidepressant, or antipsychotic), and disease-specific information (CAG, CAP).

Covariate selection was based on the posterior distribution of each covariate coefficient β_j_. Covariates were considered important if the 90% posterior distribution of their coefficient β_j_ did not include 0. Conversely, if the distribution included 0, the covariates were deemed unimportant and subsequently removed from further analysis. Covariate selection was performed independently for each clinical score.

To address the challenge of testing multiple covariates and account for the inclusion of irrelevant predictors, we employed the horseshoe prior (Carvalho et al, 2010; Zhang et al, 2022). This prior was chosen for its ability to mitigate overfitting when assessing a large number of covariates.

A weighted linear combination of the selected covariates was used as a predictor of the progression rate for each clinical score. For more detailed information, please refer to the methods section.

#### Software

Figures and statistical analysis were performed in Julia (version 1.9.0).

## Results

### Distinct trajectories of functional, motor, and cognitive decline

To determine the disease progression trajectories for each clinical score over the course of the disease, we analyzed natural history data from Enroll-HD. For that, we developed a screening method that considered multiple trajectory shapes, such as exponential, logistic, linear, and combinations thereof (for more information, please refer to the supplementary information and Figure 1A). This method was independently applied to each UHDRS clinical score (TFC, TMS, SDMT, SWR). Our analysis revealed that TFC and TMS are best characterized by logistic-like trajectories, consistent with previous analysis (Sun et al, 2020), while cognitive scores (SDMT and SWR) display an exponential-like decline (Figures 1B, 1C). These findings indicate that different clinical scores evolve at distinct rates over the course of the disease.

**Figure 1.**
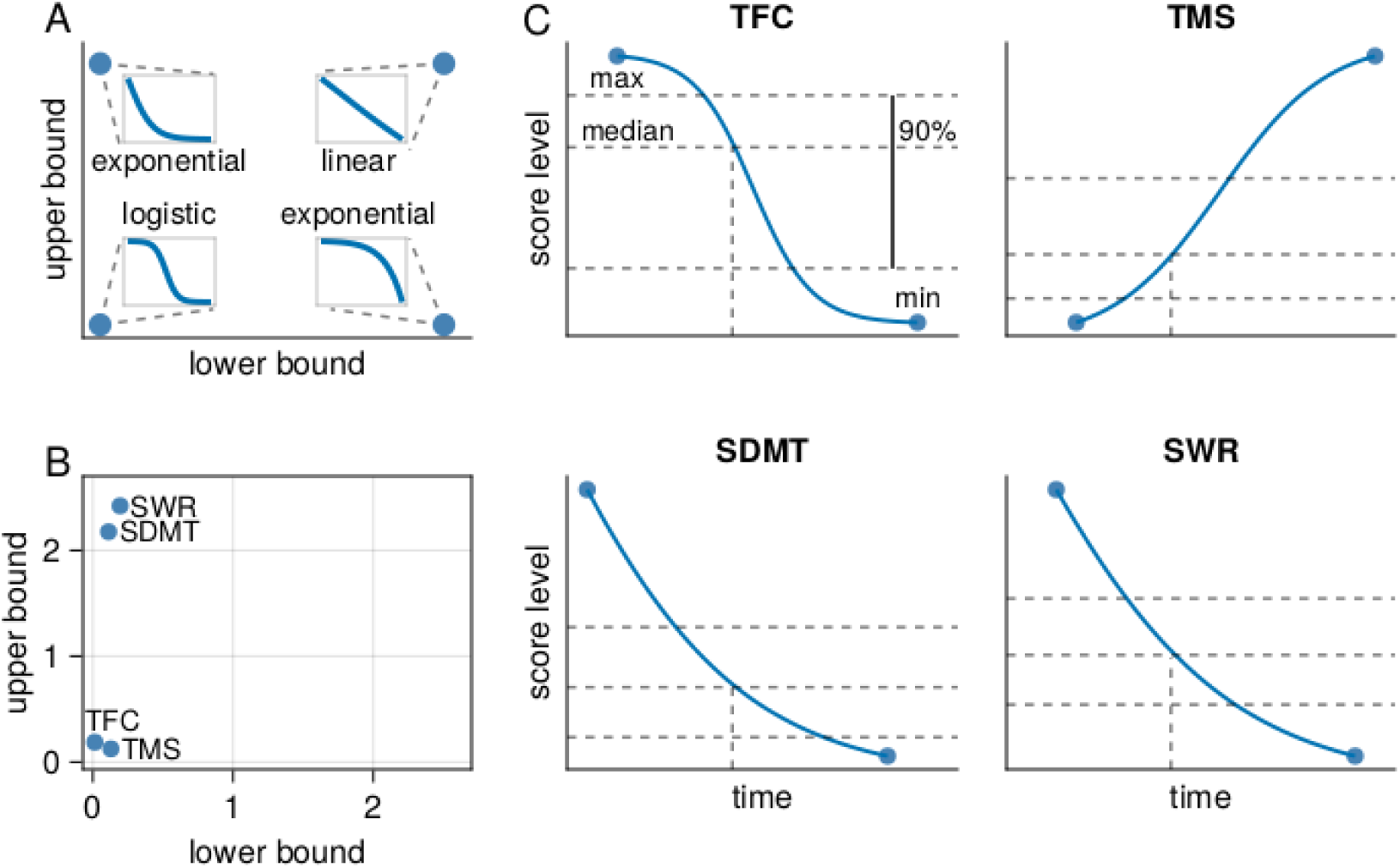
Differences in functional, motor, and cognitive trajectories. A) Schematic illustration of the disease trajectory screening procedure. By treating the lower and upper bounds of the generalized logistic model as model parameters, we screened various potential disease trajectories. Inset figures depict extreme examples: exponential, logistic, and linear models. B) TFC and TMS are best characterized by logistic-like trajectories (low lower and low upper bound values), while cognitive scores (SDMT and SWR) are best characterized by exponential-like trajectories (low lower and high upper bound values). C) Fitted trajectories for TFC, TMS, SDMT and SWR. Dots at the extreme represent minimum and maximum score levels in the data. Dashed lines represent the median and 90% interval. In contrast to other scores, TMS values increase with the progression of the disease. These trajectories provide an average representation of the expected progression over the course of the disease (time).

### Baseline characteristics as strong predictors of clinical decline

After defining disease trajectories for each clinical measurement, we further explored whether baseline patient characteristics could predict the rate at which participants from the Enroll-HD cohort progress along these predefined trajectories. Previous studies have shown that baseline score levels, CAP, CAG length, co-medication are predictive of progression rate (Sun et al, 2020, Ghazaleh et al, 2021). Based on these studies, we selected 17 patient characteristics representing disease stage, patient demographics, co-medication use, and disease-specific information as potential predictive covariates to be evaluated (Figure 1A, Methods).

The presence of strong correlation among the selected covariates (Figure Suppl 2) poses limitations when interpreting its relevance based on their coefficients. It is important to note that a low coefficient for a correlated covariate does not imply its irrelevance, as its effect might be represented by other covariates. Keeping these limitations in mind, we observed that overall the most relevant covariates include the stage of the disease (HD-ISS and baseline clinical scores), disease-specific characteristics (CAG, CAP), and the use of tetrabenazine and antipsychotics, while patient demographics (sex, height, weight, BMI, education, age) have little/no impact. These results are consistent with previous analysis (Ghazaleh et al, 2021).

In order to validate the predictive nature of these baseline characteristics on progression rates, we performed a cross-validation analysis by randomly dividing the Enroll-HD dataset into a 70-30% training-validation set and repeating this process 100 times. We used the training set to estimate the relevance of each covariate, and then categorized patients in the validation set into two groups: the 50% more likely to progress and 50% less likely to progress based on their predicted progression rates. We observed a strong separation between these two predicted populations, indicating that baseline characteristics serve as strong predictors of progression rate and can effectively identify patients who are more likely to experience disease progression (Figure 2C).

**Figure 2.**
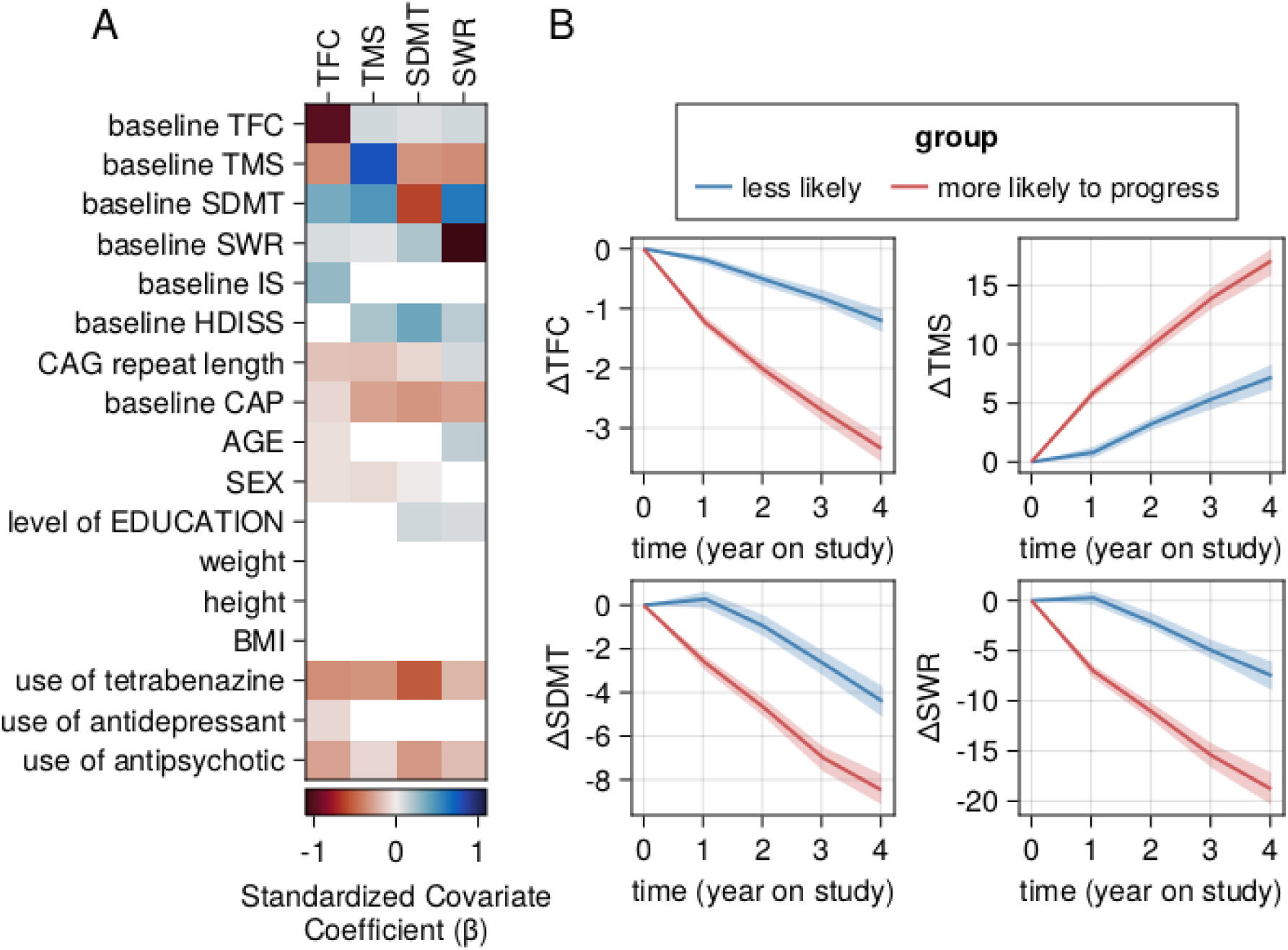
Baseline characteristics can identify patients more likely to progress. A) Heatmap illustrates the relevance of each covariate in predicting progression rate for the respective clinical scores. Darker shades indicate a stronger impact. Estimated coefficient values have been standardized according to the average progression rate to enable comparison between different clinical scores. Red shades indicate an increase in the rate of progression, while blue shades indicate a decrease. Caution should be taken when interpreting the individual effects of each coefficient due to strong correlations among the covariates (see Figure Suppl. 2). B) Cross validation analysis showing changes in TFC, TMS, SDMT, and SWR from baseline for individuals selected from Enroll-HD (validation set). Patients were categorized into 50% more likely and 50% less likely to progress based on the model’s predictions using baseline covariates. Lines represent the average changes from baseline, while the shaded area denotes a 95% confidence interval across 100 cross-validation iterations, using 70% as training and 30% as validation data from Enroll-HD.

### Weak correlation in progression rate among different clinical scores

In our initial analysis, we found that functional, motor, and cognitive scores are best characterized by distinct disease trajectories (Figure 1), implying that patients may exhibit varying rates of functional, motor, and cognitive decline throughout the course of the disease. To further examine this, we estimated the correlation of predicted progression rates among different clinical scores. Our model predicts a weak correlation in the progression rates between these clinical scores (Figure 3A). By plotting the relationship between short term changes in clinical scores from individuals selected from the Enroll-HD cohort, we further confirmed that changes in functional, motor, and cognitive scores are weakly correlated (Figure 3B). These results highlight the differential rate of progression captured by distinct clinical endpoints and emphasize that individuals who exhibit rapid progression according to one clinical score may not necessarily exhibit fast progression according to another.

**Figure 3.**
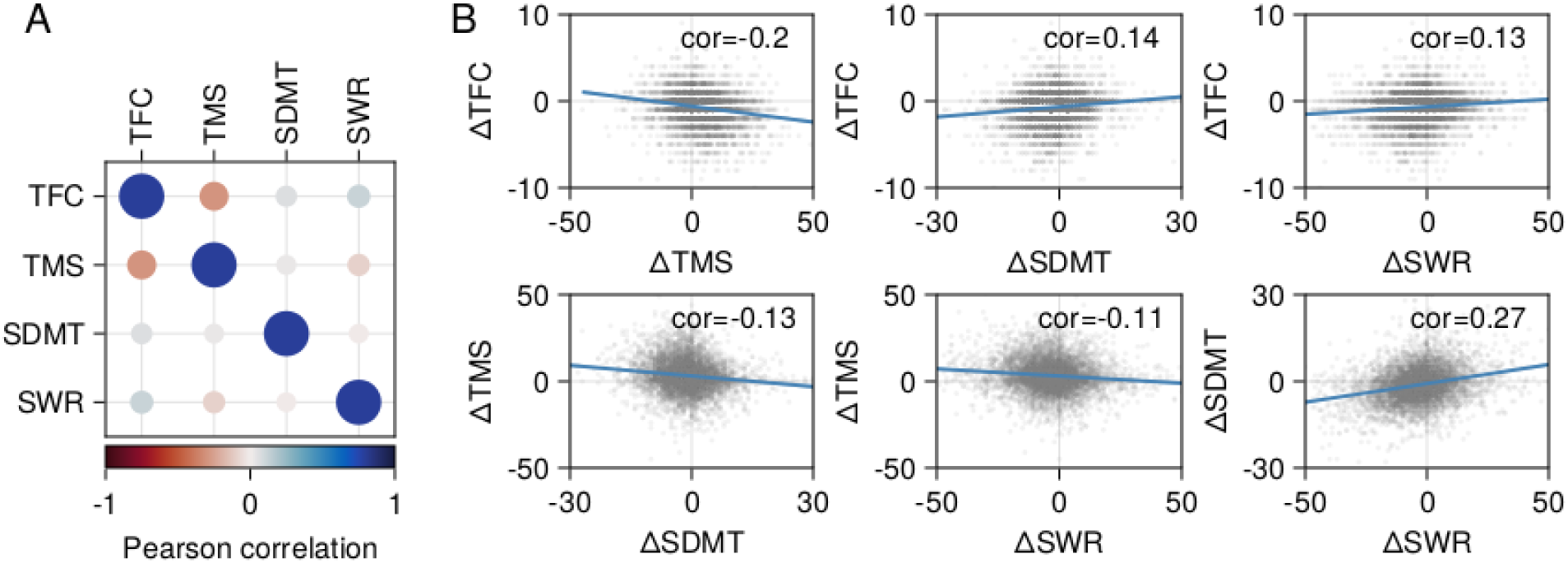
Correlation among rates of functional, motor, and cognitive decline. A) Peason correlation coefficients for the predicted progression rates among TFC, TMS, SDMT, and SWR scores. B) Associations between the one-year changes in TFC, TMS, SDMT, and SWR scores for individuals selected from the Enroll-HD database. Gray dots represent the change in each clinical score after one year for individual patients, while the blue line illustrates the linear fit of the relationship between changes in each score. Insets in the top right corner of each subplot display the correlation coefficients for the respective score changes.

### Placebo response in GENERATION HD1

Understanding and quantifying placebo response is crucial in the design of a clinical trial as it enables accurate interpretation of treatment effects and precise estimation of sample size calculations. In order to quantify the placebo response, we compared the progression in the GENERATION HD1 placebo cohort with their expected natural history progression (Figure 4A). We observed that for the functional score (TFC), the progression during the trial aligns with the natural history progression, showing no sign of a relevant placebo response. In contrast, for the motor and cognitive measurements, we found that patients in the placebo arm of GENERATION HD1 have better outcomes than expected by natural history progression (Figure 4A). We also noted that this improvement has a fast onset after treatment initiation and persists throughout the dosing period of the trial. Moreover, after this initial improvement, motor and cognitive decline follows the expected rate of decline from natural history. This finding indicates the presence of a placebo response that contributes to a noticeable improvement in motor and cognitive measurements within a few weeks following the start of the trial, and this improvement persists consistently throughout the trial duration. (Figure 4A).

**Figure 4.**
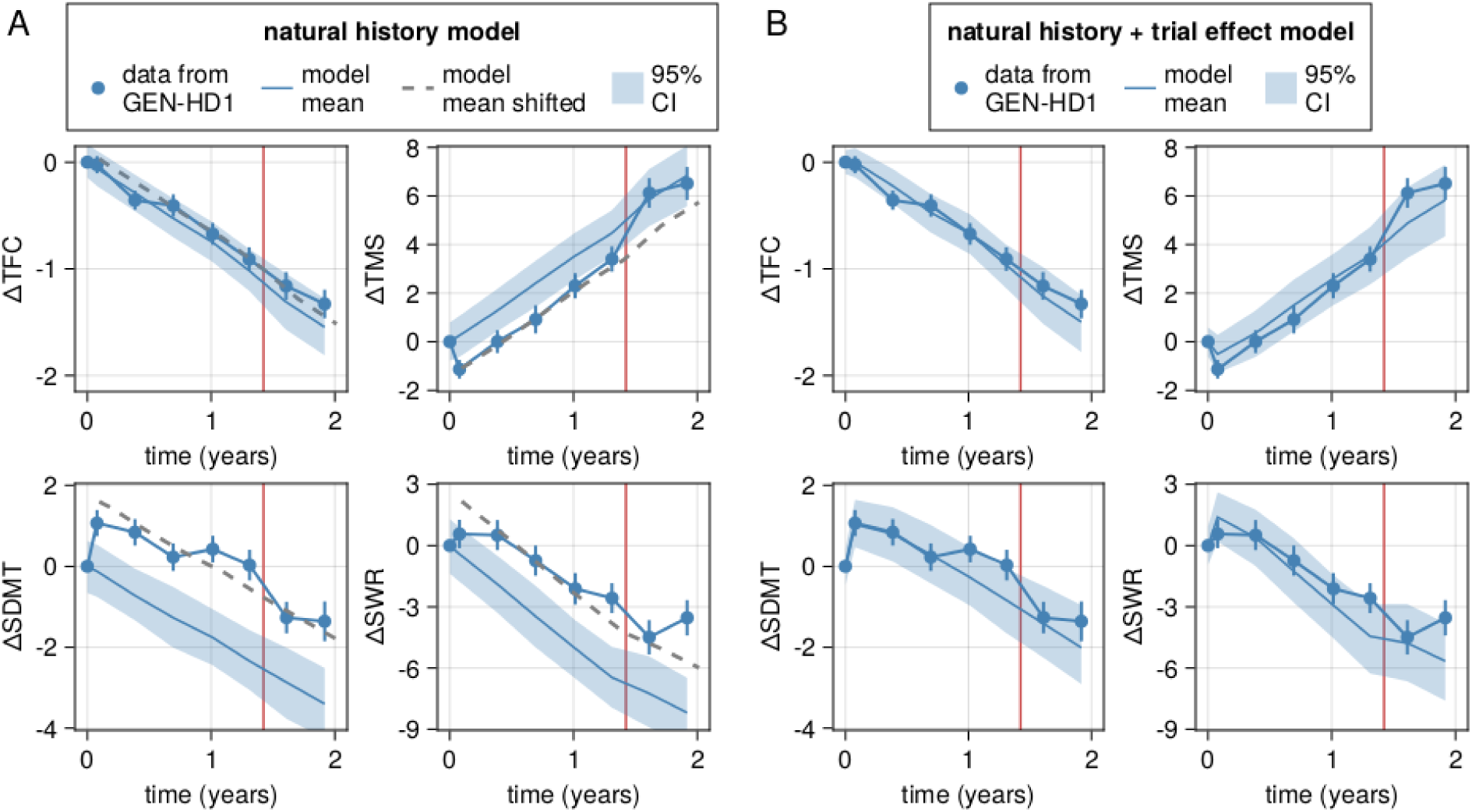
Trial effects on functional, motor, and cognitive scores. This figure illustrates the changes from baseline in TFC, TMS, SDMT, and SWR. Blue dots and lines indicate the average data from the GENERATION HD1 placebo arm, while vertical blue lines represent the standard error. The vertical red line marks the end of the dosing period for most patients (after week 69). A) The blue line shows the prediction based on the natural history model, with the shaded area representing the 95% confidence interval. The dashed line indicates a shift from the mean predictions of the model, demonstrating that by adjusting the model prediction proportionally to the initial improvement, the progression rate in GENERATION HD1 aligns with the predicted natural history. B) The blue line and shaded area depict the combined natural history and trial effect model’s average and 95% confidence interval prediction. For the composite score (cUHDRS) please refer to Figure S7.

Intriguingly, we identified differences in the placebo response between motor and cognitive scores. An offset of the response was observed for the motor score (TMS) after the end of the trial dosing period, implying the offset of a placebo effect. However, this was not the case for the cognitive measurements, where we observed no offset of this initial improvement after dosing was stopped, suggesting a different placebo response, potentially a learning or practice effect that persists after dosing was discontinued (Figure 4A). By modeling placebo response as an initial improvement after baseline and constant during the dosing period of the trial (see supplementary information), our model was able to capture the changes in clinical score in GENERATION HD1 during the dosing period of the trial (up to week 69) (Figure 4B).

### Simulation of placebo arms for patient populations with different inclusion criteria

Once natural history progression, placebo response, and baseline characteristics predictive of progression rate are quantified, it is possible to simulate the changes in each clinical score for different patient populations characterized by different baseline characteristics. In Figure 5, we present the expected changes in each clinical measurement for commonly used selection criteria in HD: CAG-Age-Product (CAP), normalized prognostic index (PIN), age, and HD-ISS (for different subpopulations see Figures Suppl 3-6).

**Figure 5.**
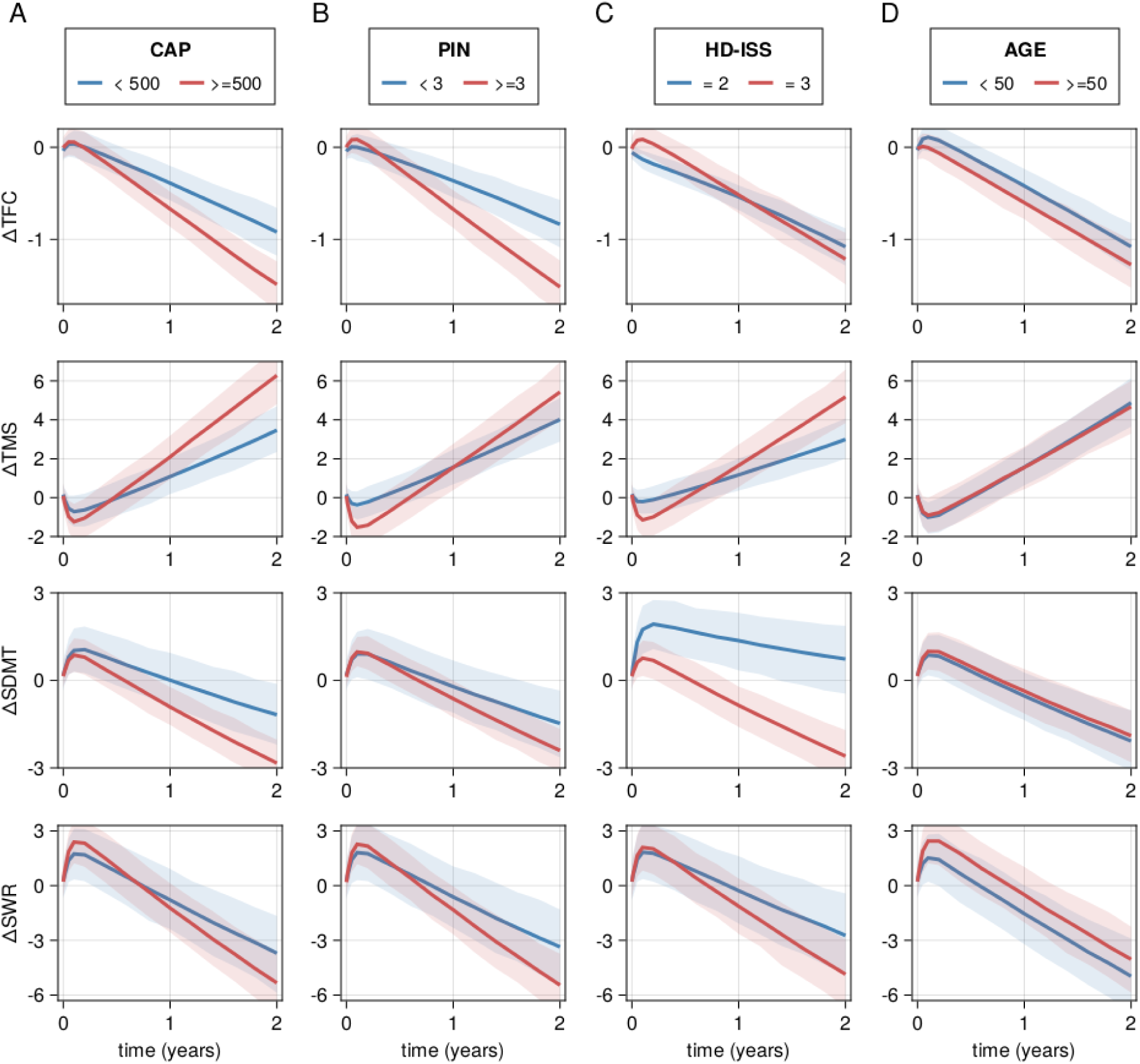
Simulation of placebo arms for different patient populations. Changes from baseline for TFC, TMS, SDMT, and SWR scores for patients segregated by A) HD-ISS, B) age, C) CAP, and D) PIN. For each scenario, we randomly sampled 250 individuals from Enroll-HD for each condition and utilized their baseline characteristics to simulate the predicted changes in the clinical score. We performed 1000 random samplings. The solid line represents the average, while the shaded area depicts the 95% confidence interval prediction. For the segregation of the plots into two groups, we chose values of CAP, PIN, and AGE that are close to the median of the Enroll-HD population (493 for CAP, 3.07 for PIN, and 51 for AGE).

We observed variations in the progression of different subpopulations depending on the specific clinical measurement utilized. For instance, PIN and CAP can effectively enrich the population with patients more likely to progress, particularly when utilizing TFC or TMS (Figure 5A,B). Moreover, when using TFC as an endpoint, the progression rate in individuals with HD-ISS = 2 is not expected to be different from those with HD-ISS = 3, in contrast to other clinical measurements (Figure 5C). Lastly, no relevant difference is observed when selecting by age (Figure 5D). These results highlight the importance for careful selection of appropriate clinical endpoints and consideration of specific subpopulations when designing and analyzing clinical trials for HD.

## Discussion

In this study, we aimed to understand and quantify disease progression and placebo response in the UHDRS clinical measurements used to assess functional (TFC), motor (TMS), and cognitive (SDMT, SWR) progression in HD. We build on previous work and the availability of large natural history cohort data (Enroll-HD) to model the natural history progression of these clinical scores. We developed a screening method to estimate disease trajectory for each clinical score and found that TFC and TMS were best characterized by logistic-like trajectories, consistent with previous analysis (Sun et al, 2020), while cognitive scores (SDMT, SWR) exhibited exponential-like trajectories. We further investigated whether baseline characteristics available in the Enroll-HD cohort could serve as predictors of the progression rates of each clinical score. Our findings indicate that baseline characteristics act as robust predictors of progression rate for each clinical score. Interestingly, when comparing the progression rates among different scores over one year, we observed a weak correlation among them, indicating that patients who are likely to experience progression in one clinical score may not necessarily exhibit the same progression in another score in the short term.

Furthermore, we sought to understand the nature and magnitude of placebo response by analyzing data from the GENERATION HD1 placebo arm. Our analysis revealed minimal placebo response in the functional score (TFC). In contrast, we observed a strong placebo response with a clear onset and offset in the motor score (TMS), suggestive of a placebo effect. Interestingly, the cognitive scores (SDMT, SWR) exhibited an even stronger placebo response without an observed offset after the dosing period of the trial ended. This suggests that the placebo response observed in cognitive scores may be related to a learning/practice effect, potentially influenced by the frequency of assessments in the GENERATION HD1 trial compared to the Enroll-HD cohort. Importantly, these effects also significantly influence the composite cUHDRS score, given that it is derived from these individual metrics.

Taken together, these findings have significant implications for trial design in HD. The strong predictive value of baseline characteristics for progression rate allows for the selection of patients based on their expected rate of progression, enabling for example trial enrichment with patients more likely to progress over the period of the trial. However, the weak correlation in progression rates between different clinical scores indicates that patients likely to progress based on one score may not necessarily progress based on other scores. This is because the clinical scores change in a non-linear manner, i.e. with different speeds at different moments of the disease. Therefore, depending on where patients are in the course of the disease, they are more likely to progress according to one endpoint but not another. This highlights the importance of carefully selecting endpoints when enriching trials with specific patient populations. Furthermore, the observed differences in the nature and magnitude of the placebo response highlight the need for a better understanding of their underlying effects. It would be critical to understand whether such a placebo response is seen in other trials and whether the magnitude of such effects are similar to those observed in GENERATION HD1. Moreover, previous research has shown that a composite measure of functional, motor, and cognitive scores (cUHDRS) provides an improved measure of clinical progression and enhances clinical trial design by requiring smaller sample sizes (Schobel et al., 2017). The use of this composite score is becoming common practice, however, because these analyses were evaluated in the absence of trial data, future work should evaluate if these conclusions hold in the presence of placebo response.

Importantly, our analysis revealed that a placebo response initially leads to an improvement after baseline, but after accounting for this initial improvement, the rate of progression between the GENERATION HD1 placebo arm and Enroll-HD cohorts remained consistent. This highlights the value of Enroll-HD as a retrospective source of natural history information.

Our mathematical framework represents an initial step towards quantifying changes in clinical endpoints within a clinical trial setting. By incorporating both natural history progression inferred from Enroll-HD and placebo response inferred from GENERATION HD1, our model effectively captures changes in clinical endpoints for different patient populations, and can be used to optimize the design and analysis of clinical trials in HD. Moreover, our model can serve as the first step towards having a modeling tool for simulating virtual placebo arms, which can optimize trials by reducing the number of required patients and accelerating clinical research.

It is important to acknowledge the main limitation of the current model, which lies in its reliance on a limited amount of trial data (260 patients from GENERATION-HD1), with the most data being up to 69 weeks. Therefore, the observed placebo response in GENERATION-HD1 may not necessarily be generalized to other trials with different patient populations and trial designs. Moreover, while Enroll-HD includes participants from various stages of the disease, it is worth noting that certain stages, such as early stages, may be less represented. If future trials focus on these specific populations, it may be necessary to adapt the model accordingly to ensure its applicability.

In summary, our findings bring a new understanding of disease progression and placebo response in HD and our theoretical framework has the potential to enhance the efficiency of clinical trials and accelerate the development of effective treatments for HD patients.

## Data Availability

Enroll-HD periodic data set 5 (PDS5) is accessible through the Enroll-HD website (enroll-hd.org/).
For clinical trial studies, qualified researchers may request access to individual patient-level clinical data through a data request platform. At the time of writing, this request platform is Vivli. https://vivli.org/ourmember/roche/. For up-to-date details on Roche's Global Policy on the Sharing of Clinical Information and how to request access to related clinical study documents, see here: https://go.roche.com/data_sharing. Anonymized records for individual patients across more than one data source external to Roche cannot, and should not, be linked due to a potential increase in risk of patient re-identification.

## Acknowledgements

Natural history data used in this work were generously provided by the participants in the Enroll-HD study and made available by CHDI Foundation, Inc. Enroll-HD is a clinical research platform and longitudinal observational study for Huntington’s disease families intended to accelerate progress towards therapeutics; it is sponsored by CHDI Foundation, a nonprofit biomedical research organization exclusively dedicated to collaboratively developing therapeutics for HD. Enroll-HD would not be possible without the vital contribution of the research participants and their families

We thank all CHDI/Enroll-HD as well as all the participants in GENERATION HD1, their families, the site staff and trial team, for their time and commitment to the trial; and the the following colleagues and HD experts for their critical feedback on method development and discussions: Francesco Brizzi, Tony Kam-Thong, Antoine Soubret, Ben Ribba, and David Hawellek.

## Supporting information

### Mathematical model

In our methodology, we begin by estimating the disease trajectories for each specific clinical score throughout the progression of the disease. To achieve this, we employ a generalized logistic model, where the lower (l) and upper (u) bounds are parameters that are inferred from the available data for each score. Subsequently, we infer the inter-individual variability in progression rate (Ω*_R_*), residual unexplained variability (σ*_NHT_*), and the relevance of each covariate *j* in predicting progression rate represented by the covariate coefficient 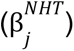. To accomplish this, we utilize data from Enroll-HD.

Parameters related to placebo response are inferred using the GENERATION HD1 placebo cohort. Similarly to the progression rate, we infer the inter-individual variability in the placebo response (Ω*_A_*), residual unexplained variability (σ*_PLB_*), and identify the relevance of each covariates in predicting the placebo response 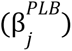.

Below we describe the details of our mathematical framework, and the estimated parameters can be found in tables S1-4.

### Natural history progression model

In order to infer the most suitable disease trajectory for each clinical score, we defined a scaling function *f* so that once it is applied to a score (*x_i_*) of individual *i*, it renders a progression rate (*r*) that is linearly changing in time, i.e:

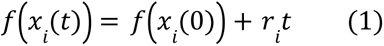

To define *f*, we used the generalized logistic model with upper (*u*) and lower (*l*) bound as parameters to be inferred. The generalized logistic model can be defined as:

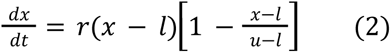

with the following analytical solution:

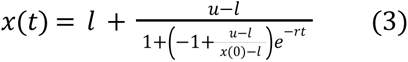

which can be rewritten as:

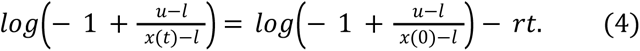

Now, by defining:

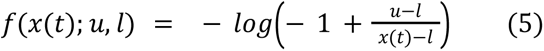

the solution becomes linear in time (as defined in Eq. 1):

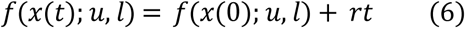

and changing the values *u*and *l* enables screening for many potential non-linear trajectories (see figure 1A).

Once *u* and *l* values are estimated, equation 5 can be used to rescale the values of the clinical score (*x*), and the model can be described as a linear model in time (Eq. 1). For simplicity, let’s define *y*(*t*) = *f*(*x*(*t*); *u*, *l*) as the scaled clinical score values, and *r_i_* as the individual progression rate. With that, changes from baseline (*t* = 0) can be described as:

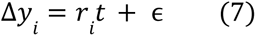

with ɛ ∼ *N*(0, σ) representing the residual unexplained variability.

In order to estimate the individual progression rate *r_i_*, we assume that

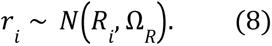

where Ω*_R_* represents the inter-individual variability in progression rate and is assumed to be constant and inferred from data. The individual average progression (*R_i_*) is assumed to be linearly dependent on predictive baseline characteristics (covariates) such that:

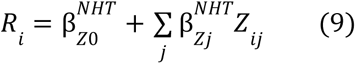

where *Z_ij_* is the z-score value of covariate *j* for individual *i* for continuous covariates and either 0 or 1 in case of binary covariates, and 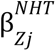 represents the weight of covariate *j* in predicting the progression rate.

To estimate the weight of each covariate 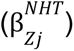 we applied HorseShoe priors. We found that R2D2 HorseShoe prior, as proposed by Zhang and colleagues (Zhang et al, 2022), proved to be effective in this context.

### Placebo response

In a clinical trial setting, the expectations of patients or clinicians regarding the potential effects of a drug can impact the measurements of clinical scores. In order to identify potential placebo responses, we compared the progression in the GENERATION HD1 placebo cohort with their expected natural history progression (Figure 4A). We found that placebo response can be characterized by an improvement after baseline that appears to remain constant throughout the dosing period of the trial (Figure 4A). We observe this for all clinical scores. Based on these observations, we represented the placebo response mathematically as a quick improvement after baseline (t=0). The parameters related to progression rate are inferred using Enroll-HD, and considered as informative priors when analyzing clinical trial data. Therefore, for the placebo arm, the model becomes:

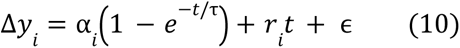

with

τ = 0. 05 (<< 1 year) (i.e. rapid improvement after baseline and constant afterwards). The parameter α*_i_* represents the individual trial effect which is assumed to be normally distributed:

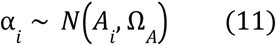

where Ω*_A_* represents the inter-individual variability and the average *A_i_* is assumed to be linearly dependent on baseline characteristics (covariates) :

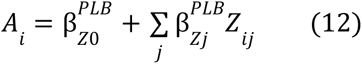

where *Z_ij_* is the z-score value of covariate *j* for individual *i* for continuous covariates and either 0 or 1 in case of binary covariates, and 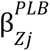 represents the weight of covariate *j* in predicting the progression rate. To estimate the weight of each covariate we applied HorseShoe priors.

**Model Fitting and Diagnostics:** The model was fit using a Markov Chain Monte Carlo (MCMC) approach with 5 chains of 200 iterations each, discarding the first 1000 iterations as burn-in. Convergence was evaluated using the potential scale reduction factor (R-hat) and the model fit was assessed through visual predictive checks (refer to Figure S3-6, S8).

#### Parameters

In our analysis, we used the z-transformed value of the covariates to determine the covariate coefficients (Eq 9). For that, each covariate *j* for each individual *i*, is transformed according to the following equation:

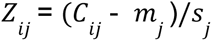

where *m_j_* and *s_j_* are the mean and standard deviation of the continuous covariates *C_ij_* at Enroll-HD. For binary covariates, we assumed *m_j_* = 0 and *s_j_* = 1.

In order to avoid having to make this transformation in future analysis, we can rewrite equation 9 as:

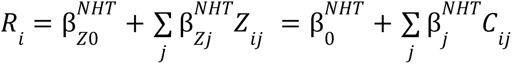

where

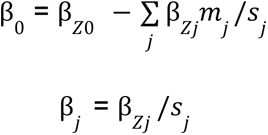

And the values at table S2 represent the values of β*_j_* . With that the estimated progression rate for each patient can be inferred based on their baseline characteristics.

## Author Contributions

M.B., Y.Y, P.M, C.D, and P.S.D. contributed to the conception and design of the study; M.B. contributed to the acquisition and analysis of data; M.B., Y.Y, J.D.L, C.S., P.M, C.D, and P.S.D. contributed to drafting the text or preparing the figures.

## Funding

Supported by F. Hoffmann–La Roche.

## Data Sharing Statement

Enroll-HD periodic data set 5 (PDS5) is accessible through the Enroll-HD website (enroll-hd.org/).

For clinical trial studies, qualified researchers may request access to individual patient-level clinical data through a data request platform. At the time of writing, this request platform is Vivli. https://vivli.org/ourmember/roche/. For up-to-date details on Roche’s Global Policy on the Sharing of Clinical Information and how to request access to related clinical study documents, see here: https://go.roche.com/data_sharing. Anonymized records for individual patients across more than one data source external to Roche cannot, and should not, be linked due to a potential increase in risk of patient re-identification.

---

**Figure S1.**
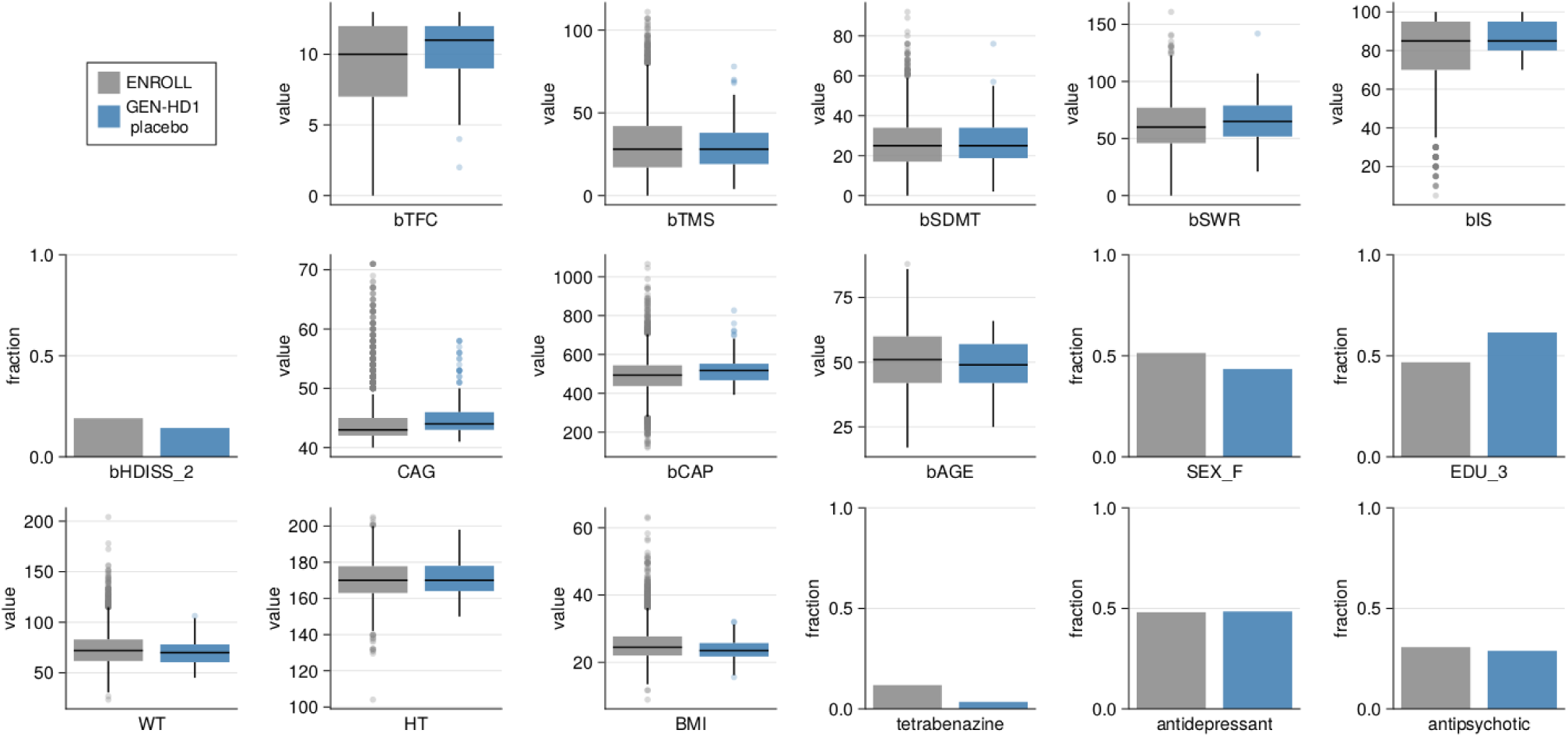
Comparison of baseline patients characteristics between Enroll-HD and GENERATION-HD1. The term EDU_3 refers to an education level (EDU) greater than 3, bHDISS_2 denotes a baseline HDISS equal to 2, and SEX_F denotes sex equal to female.

**Figure S2.**
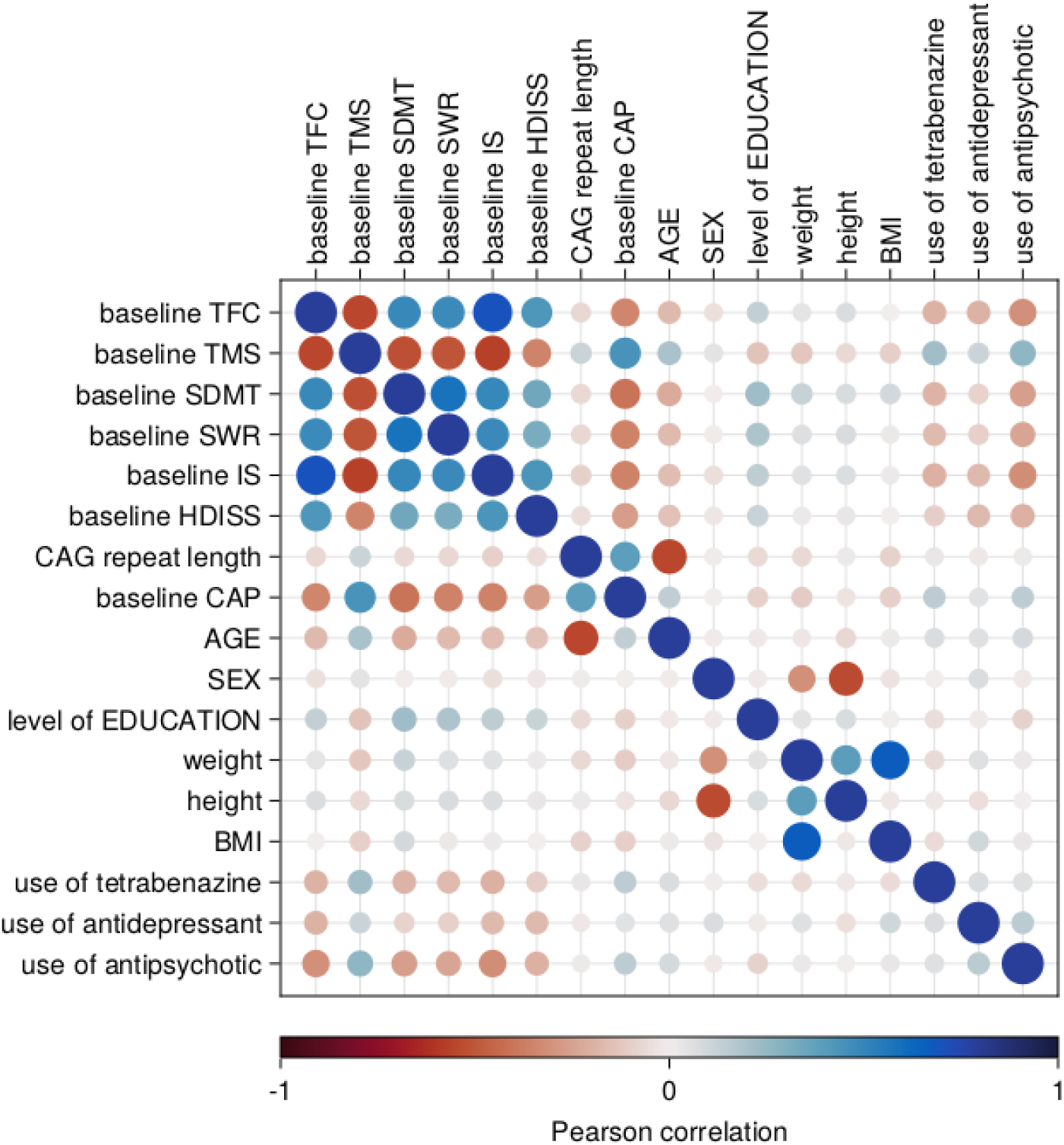
Pearson correlation coefficients among the covariates.

**Figure S3.**
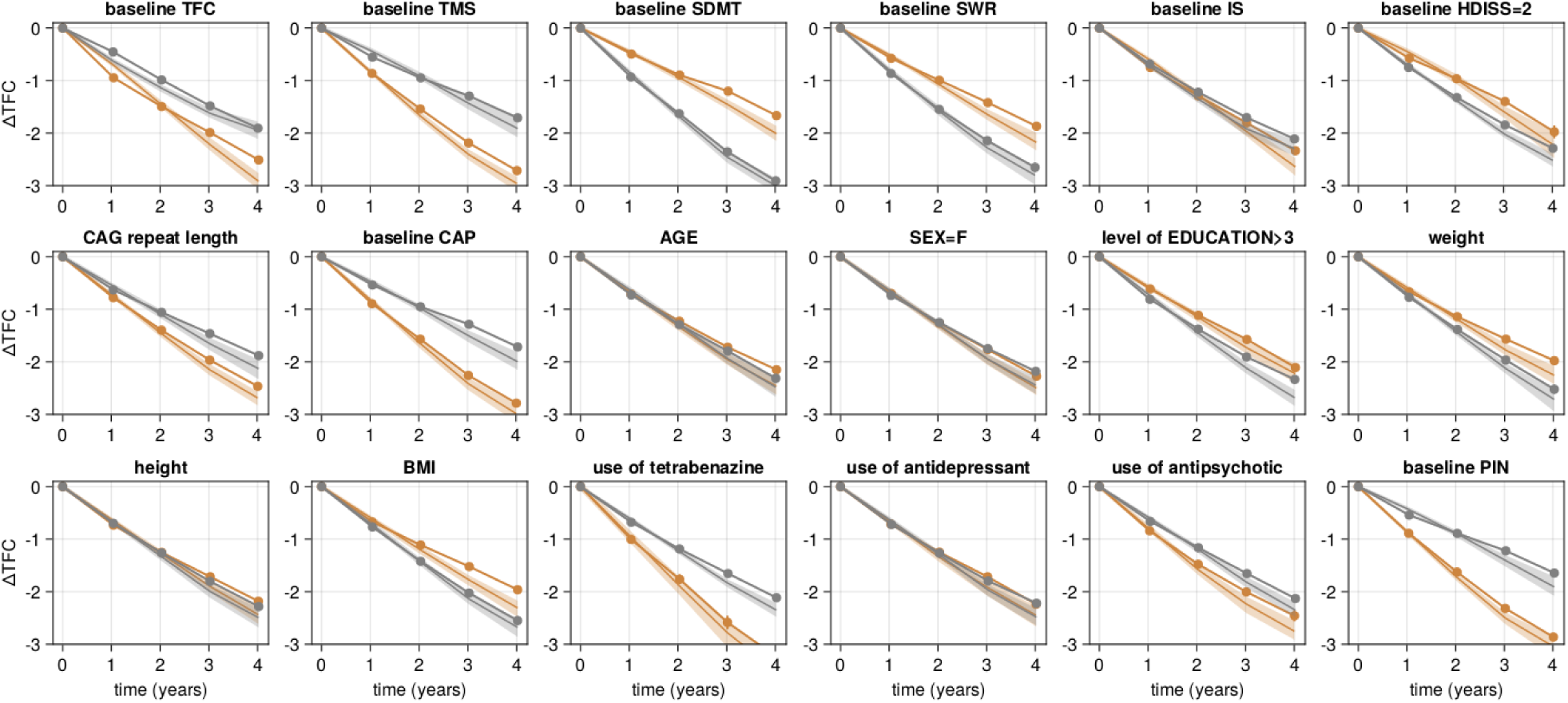
Visual Predictive Check for different patient subpopulations. The model’s visual predictive checks demonstrate a high degree of consistency with the data when segmented into subpopulations. The dots symbolize the average data derived from Enroll-HD, while the lines depict the model’s average predictions. The encompassed area signifies the 90% confidence interval (CI). Participants were categorized based on their baseline covariates, as indicated in the title. For continuous covariates, a median split was employed, with gray signifying values lower than the median and brown indicating values higher than the median. For discrete covariates, gray represents false, and brown signifies true.

**Figure S4.**
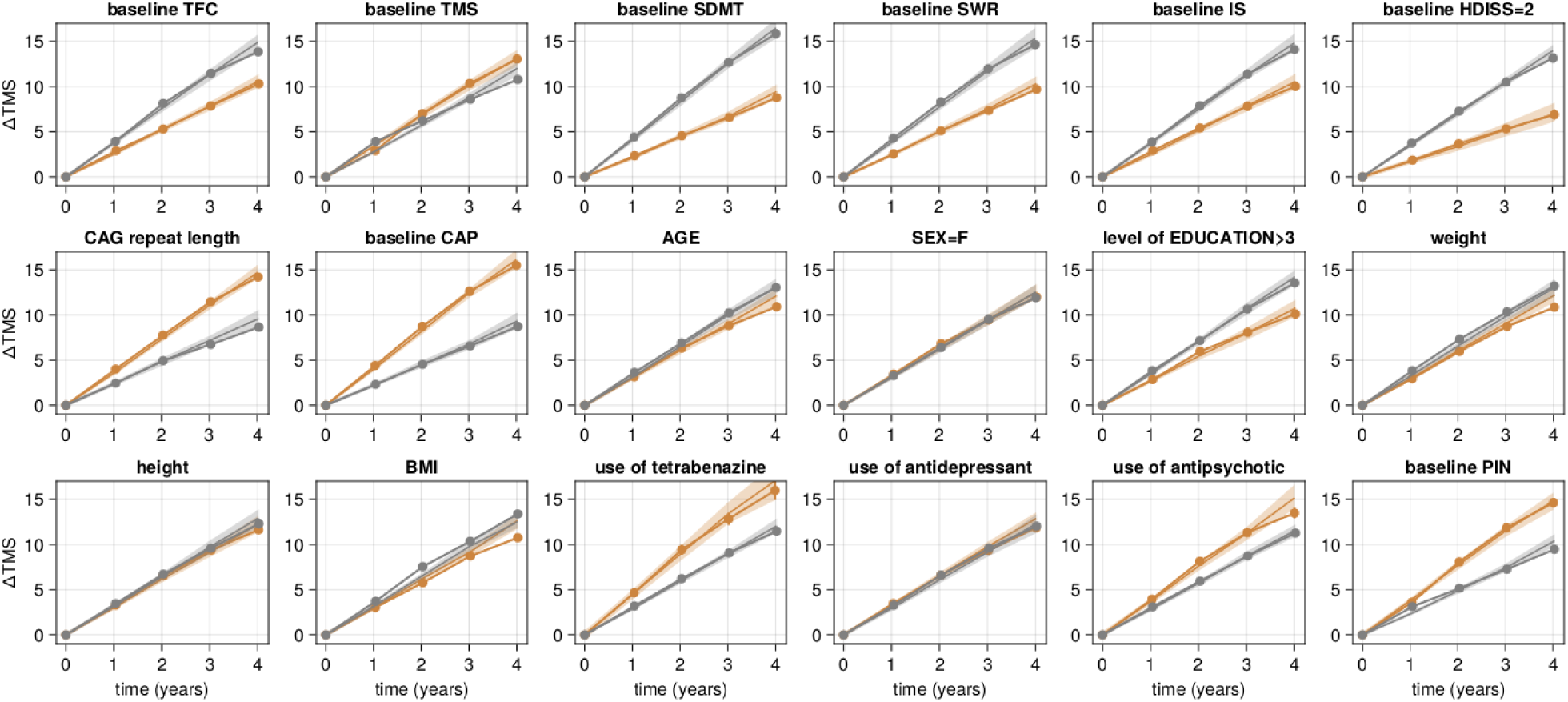
Visual Predictive Check for different patient subpopulations. Same as figure S3 for TMS score.

**Figure S5.**
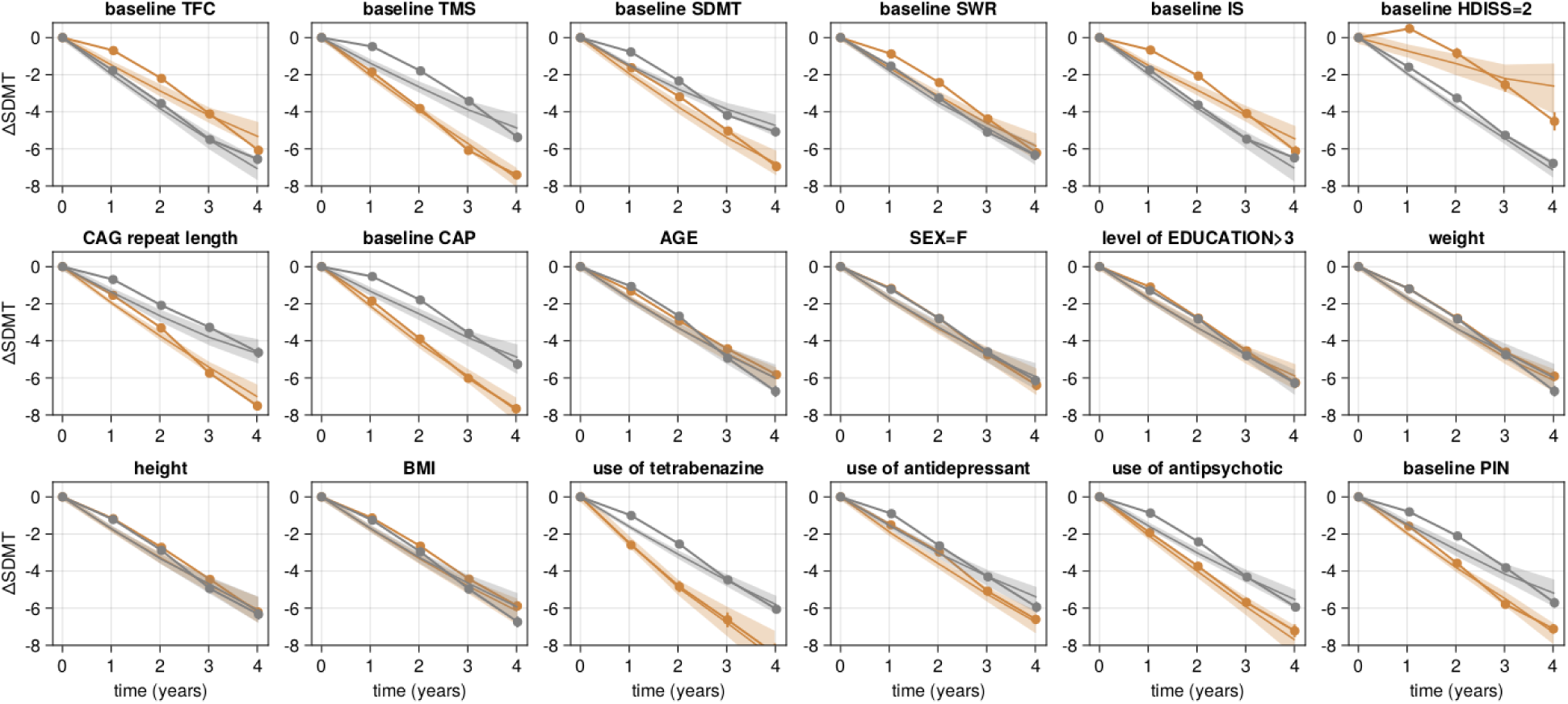
Visual Predictive Check for different patient subpopulations. Same as figure S3 for SDMT score.

**Figure S6.**
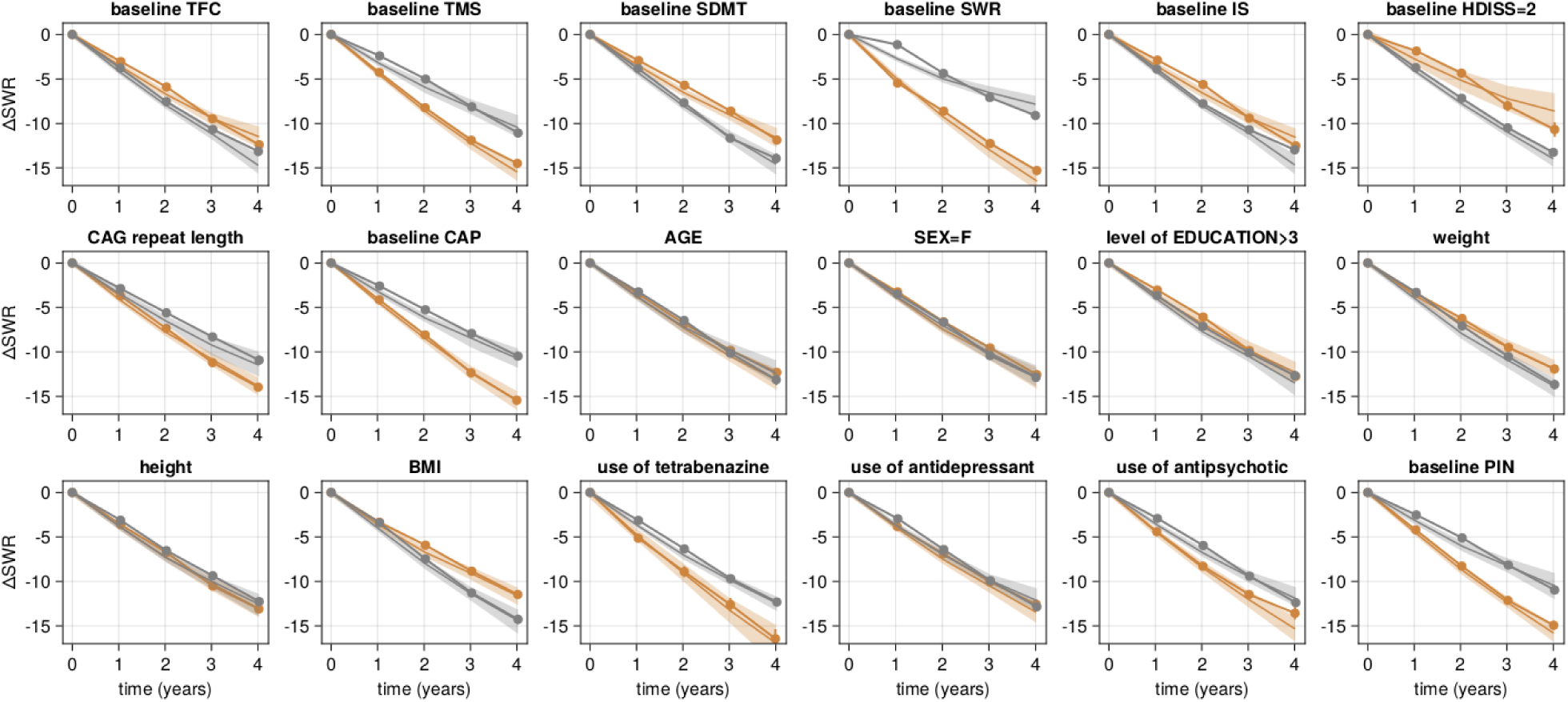
Visual Predictive Check for different patient subpopulations. Same as figure S3 for SWR score.

**Figure S7.**
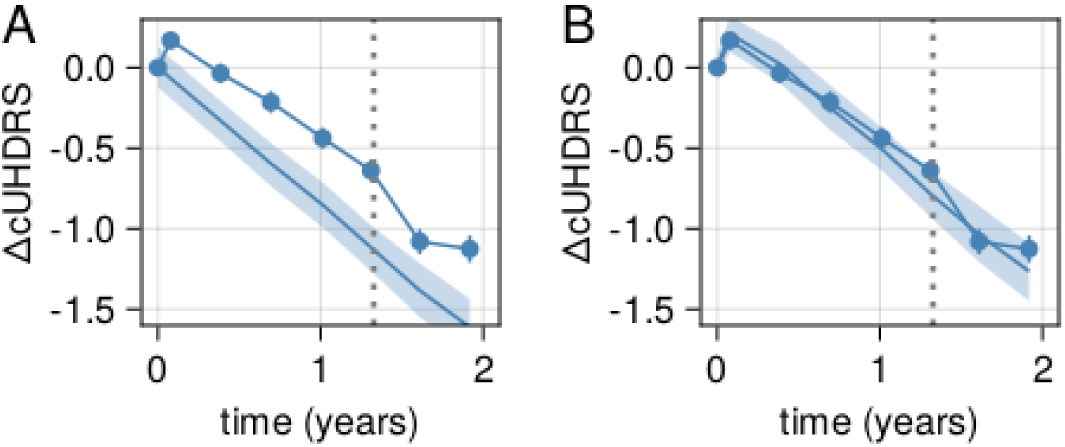
Placebo response on composite score (cUHDRS). Same as Figure 4 for the composite cUHDRS.

**Figure S8.**
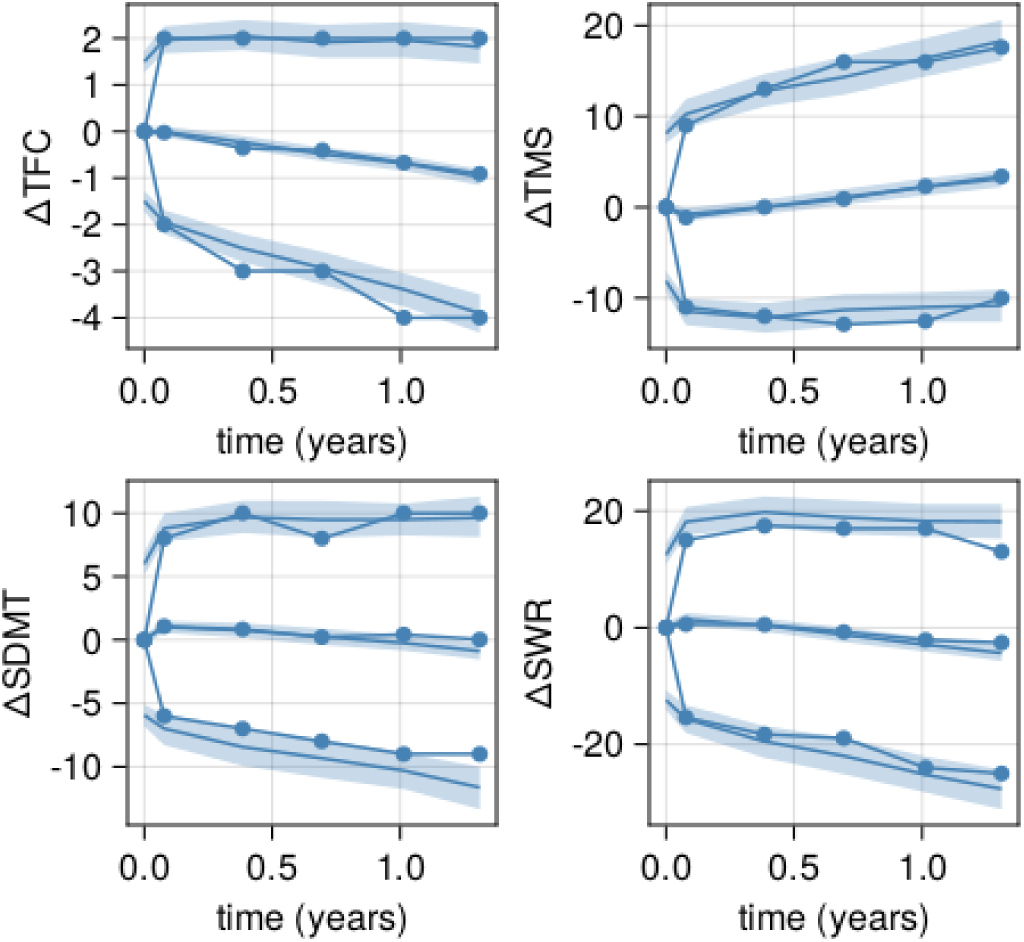
5th and 95th percentile VPC. Visual predictive checks for average, 5th and 95th percentile changes from baseline.

**Table S1:** Parameters related to trajectory screening. The parameters xmin and xmax represent the minimum and maximum values in Enroll-HD for each clinical score. The parameters u and l represent the parameters used in Eq 2, and are estimated as l = xmin - lower*(xmax - xmin) and u = xmax + upper*(xmax-xmin). The upper and lower parameters represent fractions of the total score range added to the data limits (xmin or xmax). This approach facilitates convenient comparison between different endpoints, as illustrated in Figure 1A.

| u/l bounds | TFC | TMS | SDMT | SWR |
| --- | --- | --- | --- | --- |
| xmin | 0.00 | 0.00 | 0.00 | 0.00 |
| xmax | 13.00 | 120.00 | 104.00 | 161.00 |
| l | -0.19 | -15.51 | -11.69 | -31.42 |
| u | 15.42 | 134.95 | 330.21 | 550.68 |
| lower | 0.01 | 0.13 | 0.11 | 0.20 |
| upper | 0.19 | 0.12 | 2.18 | 2.42 |

**Table S2:**
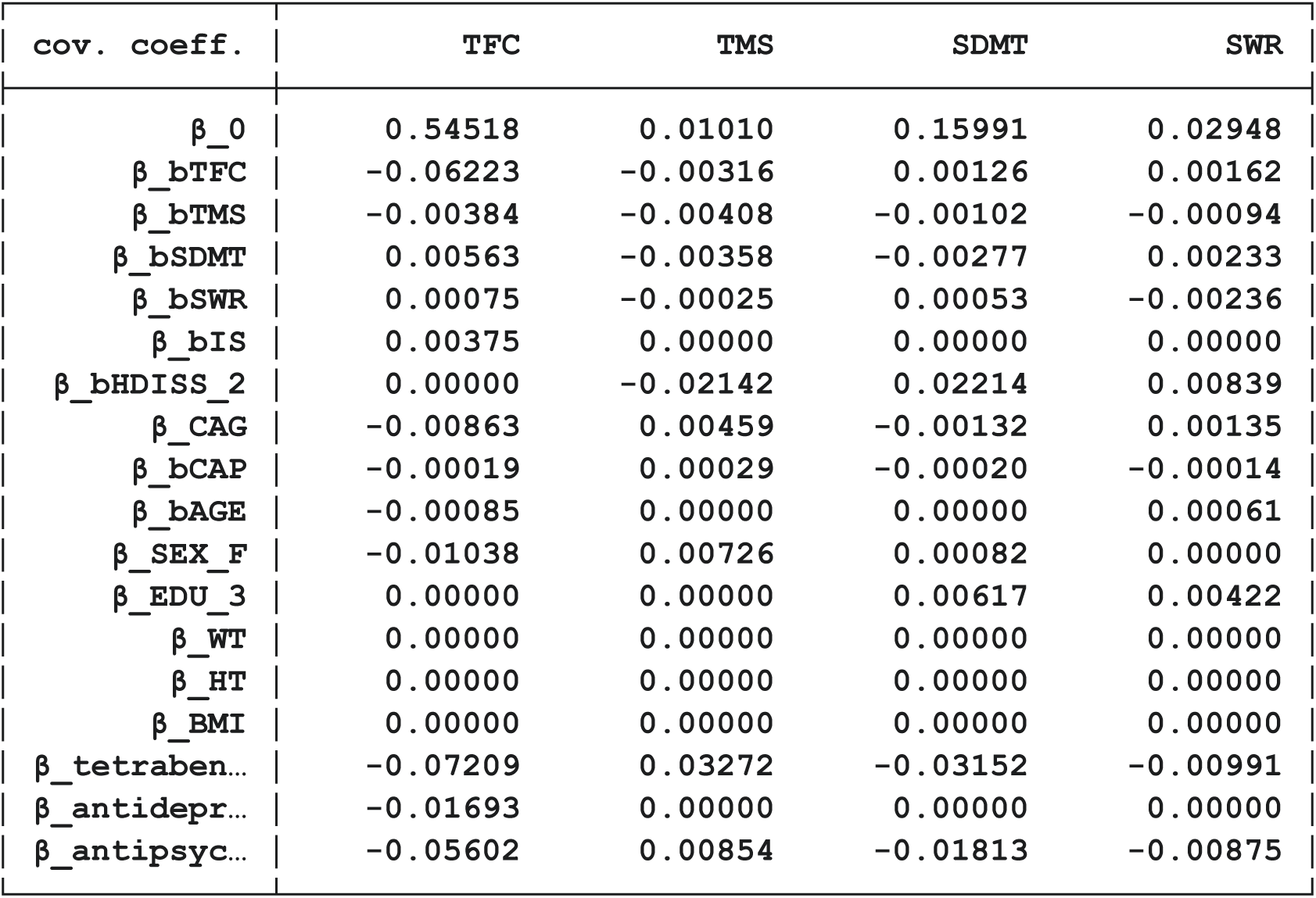
Covariate coefficients predictive of progression rate.

**Table S3.**
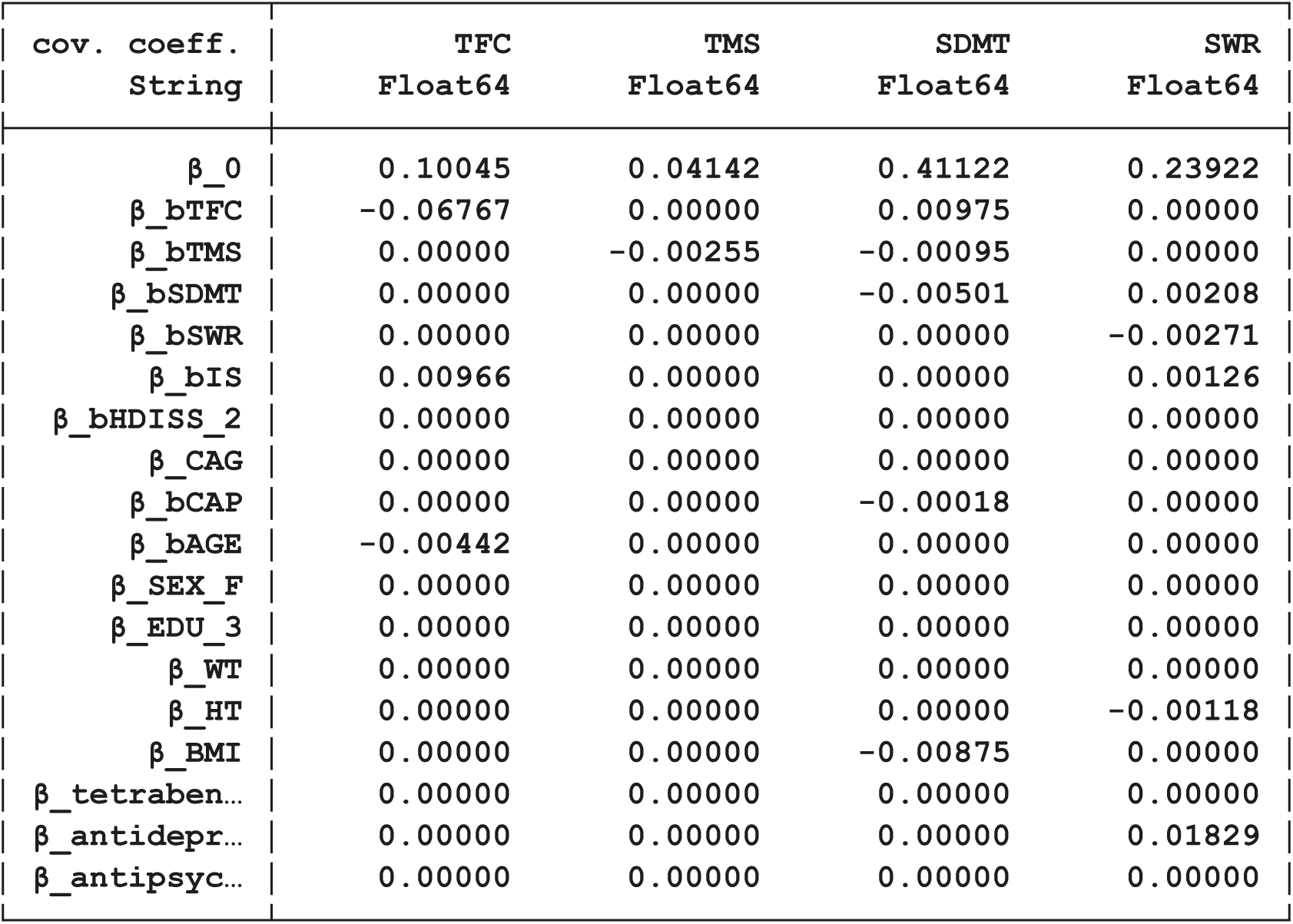
Covariate coefficients predictive of placebo response.

**Table S4.** Standard deviations for natural history and placebo arm model.

| std param. | TFC | TMS | SDMT | SWR |
| --- | --- | --- | --- | --- |
| $\Omega_R$ | 0.20254 | 0.09818 | 0.06706 | 0.05330 |
| $\sigma_{NHT}$ | 0.42798 | 0.22428 | 0.15593 | 0.13635 |
| $\Omega_A$ | 0.26841 | 0.18748 | 0.10695 | 0.10043 |
| $\sigma_{PLB}$ | 0.29052 | 0.16403 | 0.10791 | 0.09387 |

## Notes

### Competing Interest Statement

The authors have declared no competing interest.

### Author Declarations

Enroll-HD periodic data set 5 (PDS5) is accessible through the Enroll-HD website (enroll-hd.org/). For clinical trial studies, qualified researchers may request access to individual patient-level clinical data through a data request platform. At the time of writing, this request platform is Vivli. https://vivli.org/ourmember/roche/. For up-to-date details on Roche's Global Policy on the Sharing of Clinical Information and how to request access to related clinical study documents, see here: https://go.roche.com/data_sharing. Anonymized records for individual patients across more than one data source external to Roche cannot, and should not, be linked due to a potential increase in risk of patient re-identification.

## References

Abeyasinghe, P. M., Tabrizi, S. J., Scahill, R. I., Durr, A., Roos, R. A., Leavitt, B. R., … & Stout, J. C. (2021). Tracking Huntington’s disease progression using motor, functional, cognitive, and imaging markers. Movement Disorders, 36(10), 2282–2292.

Andrew, S. E., Paul Goldberg, Y., Kremer, B., Telenius, H., Theilmann, J., Adam, S., … & Hayden, M. R. (1993). The relationship between trinucleotide (CAG) repeat length and clinical features of Huntington’s disease. Nature genetics, 4(4), 398–403.

Byrne, L. M., et al (2017). Neurofilament light protein in blood as a potential biomarker of neurodegeneration in Huntington’s disease: a retrospective cohort analysis. The Lancet Neurology, 16(8), 601–609.

Carvalho, C. M., Polson, N. G., & Scott, J. G. (2010). The horseshoe estimator for sparse signals. Biometrika, 97(2), 465–480.

Ge, H., Xu, K., & Ghahramani, Z. (2018). Turing: a language for flexible probabilistic inference. In International conference on artificial intelligence and statistics (pp. 1682–1690). PMLR.

Ghazaleh, N., Houghton, R., Palermo, G., et al. (2021). Ranking the predictive power of clinical and biological features associated with disease progression in Huntington’s disease. Frontiers in Neurology.

Huntington Study Group. Unified Huntington’s Disease Rating Scale: reliability and consistency. Mov Disord. 1996;11(2):136–42.

Ko, J., Furby, H., Ma, X., et al. (2022). Clustering and prediction of disease progression trajectories in Huntington’s disease: An analysis of Enroll-HD data using a machine learning approach. Frontiers in Neurology.

Landwehrmeyer, G. B., Fitzer-Attas, C. J., Giuliano, J. D., Gonçalves, N., Anderson, K. E., Cardoso, F., … & Squitieri, F. (2017). Data analytics from Enroll-HD, a global clinical research platform for Huntington’s disease. Movement Disorders Clinical Practice, 4(2), 212–224.

Langbehn, D. R., Fine, E. M., Meier, A., & Hersch, S. (2022). Utility of the Huntington’s Disease Prognostic Index Score for a Perimanifest Clinical Trial. Movement disorders, 37(5), 1040–1046.

Lee, J. M., Huang, Y., Orth, M., Gillis, T., Siciliano, J., Hong, E., … & Gusella, J. F. (2022). Genetic modifiers of Huntington disease differentially influence motor and cognitive domains. The American Journal of Human Genetics, 109(5), 885–899.

Long, J. D., Mills, J. A., Leavitt, B. R., Durr, A., Roos, R. A., Stout, J. C., … & Landwehrmeyer, G. B. (2017). Validation of a prognostic index for Huntington’s disease. Movement Disorders, 32(2), 256–263.

McColgan, P., Thobhani, A., Boak, L., et al (2023). Tominersen in Adults with Manifest Huntington’s Disease. The New England journal of medicine, 389(23), 2203–2205.

Mohan, A., Sun, Z., Ghosh, S., et al. (2022). A machine-learning derived Huntington’s disease progression model: insights for clinical trial design. Movement Disorders.

Paulsen, J.S., Long, J.D., Ross, C.A., et al. (2014). Prediction of manifest Huntington’s disease with clinical and imaging measures: a prospective observational study. The Lancet Neurology.

Rodrigues, F. B., et al. (2020). Mutant huntingtin and neurofilament light have distinct longitudinal dynamics in Huntington’s disease. Science translational medicine, 12(574), eabc2888.

Schobel, S.A., Palermo, G., Auinger, P., et al. (2017). Motor, cognitive, and functional declines contribute to a single progressive factor in early HD. Neurology.

Sun, Z., Ghosh, S., Li, Y., et al. (2019). A probabilistic disease progression modeling approach and its application to integrated Huntington’s disease observational data. JAMIA Open.

Sun, W., Zhou, D., Warner, J.H., et al. (2020). Huntington’s disease progression: A population modeling approach to characterization using clinical rating scales. The Journal of Clinical Pharmacology.

Tabrizi, S.J., Schobel, S., Gantman, E.C., et al. (2022). A biological classification of Huntington’s disease: the Integrated Staging System. The Lancet Neurology.

Tabrizi, S. J., Scahill, R. I., Owen, G., Durr, A., Leavitt, B. R., Roos, R. A., … & Langbehn, D. R. (2012). Potential endpoints for clinical trials in premanifest and early Huntington’s disease in the TRACK-HD study: analysis of 24 month observational data. The Lancet Neurology, 11(1), 42–53.

Warner, J. H., et al. (2022). Standardizing the CAP Score in Huntington’s Disease by Predicting Age-at-Onset. Journal of Huntington’s Disease, 11(2), 153–171.

Zhang, Y., Long, J.D., Mills, J.A., et al. (2011). Indexing disease progression at study entry with individuals at-risk for Huntington disease. American Journal of Medical Genetics Part B: Neuropsychiatric Genetics.

Zhang, Y. D., Naughton, B. P., Bondell, H. D., & Reich, B. J. (2022). Bayesian regression using a prior on the model fit: The r2-d2 shrinkage prior. Journal of the American Statistical Association, 117(538), 862–874.

